# Environmental Exposures and the Human Gut Resistome in Northwest Ecuador

**DOI:** 10.1101/2025.05.23.25327954

**Authors:** Irmarie Cotto, Ana Durán-Viseras, Kelsey J. Jesser, Nicolette A. Zhou, Caitlin Hemlock, Viviana Albán, April M. Ballard, Christine S. Fagnant-Sperati, Gwenyth O. Lee, Janet K. Hatt, Charlotte Royer, Joseph N. S. Eisenberg, Gabriel Trueba, Konstantinos T. Konstantinidis, Karen Levy, Erica R. Fuhrmeister, ECoMiD Authorship Group

## Abstract

Inadequate water, sanitation, and hygiene (WASH) infrastructure may increase exposure to antimicrobial resistance (AMR). In addition, close human-animal interactions and unregulated antibiotic use in livestock facilitate the spread of resistant bacteria. We used metagenomic sequence data and multivariate models to assess how animal exposure and WASH conditions affect the gut resistome and microbiome in 53 pregnant women and 84 children in Ecuador. *Escherichia coli*, *Klebsiella pneumoniae,* and clinically relevant antimicrobial resistance genes (ARGs) were detected across all age groups, but the highest abundance was found in children compared to mothers. In mothers, higher animal exposure trended towards a higher number of unique ARGs compared to low animal exposure (β= -5.58 [95% CI: -11.46, 0.29]) and was significantly associated with greater taxonomic diversity (β= -1.29 [-1.96, -0.63]). In addition, mothers with sewer systems or septic tanks and piped drinking water had fewer unique ARGs (β= -3.52 [-6.74, -0.30]) compared to those without, and mothers with longer duration of drinking water access had lower total ARG abundance (β= -0.05 [-0.1, -0.01]). In contrast, few associations were observed in children, likely due to the dynamic nature of the gut microbiome during early childhood. Improving WASH infrastructure and managing animal exposure may be important in reducing AMR but could also reduce taxonomic diversity in the gut.

## 1.0 Introduction

Antimicrobial resistance (AMR) is a significant global public health issue, particularly affecting low, lower-middle, and upper-middle -income countries (LMICs).^1,2^ Inadequate water, sanitation, and hygiene (WASH) services, restricted access to healthcare resources, and lack of regulatory frameworks for antimicrobial use contribute to the amplified burden of AMR in these regions.^3^ Inconsistent access to clean water and adequate sanitation infrastructure can lead to increased exposure to antimicrobial residues, antimicrobial-resistant bacteria, and antimicrobial resistance genes (ARGs). Inadequate WASH also leads to an increased infectious disease burden and higher demand for antimicrobials.^4^ The World Health Organization (WHO) has emphasized the urgent need for comprehensive strategies to address AMR, noting disproportionate impacts in LMICs.^5^

Close contact with animals for household food production or income, as pets or strays, and in markets can increase microbial exchange between humans and animals in LMICs.^6–8^ Extensive use of antibiotics in food animals contributes to the development and spread of resistant bacteria and ARGs.^9^ Research assessing global trends in antimicrobial resistance has found a high prevalence of ARGs in livestock, particularly those conferring resistance to tetracyclines and beta-lactams^10^, and studies have also identified ARGs in pets.^11–13^ In LMICs, the use of antibiotics in livestock is often less regulated compared to high-income countries (HICs), leading to higher rates of AMR.^10,14^ Additionally, the burden of AMR in animals may be influenced by limited access to veterinary care.^1^ Studies in rural Ecuador have documented a diverse array of ARGs in small-scale poultry farms^15–17^ and domestic dogs^18–20^, with a high prevalence of extended-spectrum beta-lactamase (ESBL) and *mcr-1* producing *E. coli.*^21,22^ Also, research has shown a dynamic collection of horizontally transferred ARGs and mobile genetic elements (MGEs) shared between the microbiomes of children and domestic animals (i.e., dogs and chickens) in semirural Ecuadorian communities,^23^ alongside detection of enteric pathogens in both groups.^24^ While existing studies show an overall trend, location-specific decoupling of WASH characteristics and their impact on AMR is needed to implement site-specific interventions. There is also a need to understand the interplay between animal exposure and WASH conditions and AMR.

Young children are particularly at risk of exposure and vulnerable to the impacts of AMR. Behaviors such as mouthing objects and playing on the ground increase their risk of exposure to microorganisms, including those carrying antimicrobial resistance elements.^25^ Moreover, early childhood is a critical period for microbiome development, significantly influencing growth, cognitive function, and immune response.^26^ Women, especially in LMICs, also face a high risk of AMR due to biological factors like menstruation, pregnancy, and childbirth, which increase susceptibility to infections and may require antibiotic treatments.^27^ The use of unsafe sanitary products and poor sanitation during menstruation can increase infection risks, particularly urinary tract infections.^28^ During pregnancy, the need for antibiotics may be increased due to limited access to maternal healthcare and complications during childbirth.^29^ Additionally, traditional caregiving roles (e.g., healthcare work, child and elderly care and food preparation) expose women to pathogens, further elevating their risk of infection and subsequent antibiotic use.^27,30^

A group of highly virulent pathogens of public health importance, *Enterococcus faecium, Staphylococcus aureus, Klebsiella pneumoniae, Acinetobacter baumannii, Pseudomonas aeruginosa, Enterobacter spp., and Escherichia coli* (ESKAPEE pathogens) can develop resistance from ARGs circulating in the environment.^31^ The genome plasticity of these bacteria facilitates the acquisition and dissemination of ARGs through horizontal gene transfer mechanisms, often mediated by mobile genetic elements like plasmids and transposons.^32^ This adaptability enables ESKAPEE pathogens to rapidly develop resistance to multiple antibiotics, complicating treatment options and contributing to the global health threat posed by antimicrobial resistance. For instance, studies have identified the presence of various β-lactamase genes (e.g., *bla*_TEM_, *bla*_SHV_, *bla*_KPC_) in *K. pneumoniae* and other Gram-negative bacteria.^33^ Understanding the relationship between ARGs and ESKAPEE pathogens is crucial for developing effective surveillance, prevention, and treatment strategies to combat the escalating challenge of antimicrobial resistance.

While ESKAPEE pathogens are, to varying degrees, culturable, many other bacteria that harbor ARGs and could transfer them to ESKAPEE pathogens are not cultivable.^34,35^ Many investigations of AMR focus on traditional culture techniques^36^, which do not capture the full breadth of microbial diversity. Culture-based methods may neglect a significant portion of microbial diversity, resulting in an incomplete understanding of microbial communities and their associated AMR potential.^37,38^ By directly extracting and sequencing genetic material, metagenomics allows for comprehensive profiling of microbial communities, including unculturable bacteria.^39^ This approach enables the identification of novel or previously undetected ARGs, providing a more complete description of the resistome (i.e., the collection of all ARGs present in both pathogenic and non-pathogenic microbes within a given environment) beyond clinically recognized pathogens.^40^

Despite growing recognition of the role that environmental factors play in AMR, most existing studies evaluate WASH and animal exposure as broad, composite indicators and rely heavily on culture-based methods or targeted gene detection. Additionally, few studies have assessed the interactive effects of individual WASH components and animal exposure on the gut resistome in vulnerable populations such as pregnant women and young children. Our study addresses this gap by using shotgun metagenomic sequencing to investigate how specific conditions (e.g., sanitation type, drinking water availability, and animal exposure) individually and jointly shape the resistome and microbiome in a low-resource setting in Northwest Ecuador.

The objectives of this study were to (1) investigate the association of environmental factors, specifically, animal exposure, drinking water availability, drinking water source type, and sanitation systems on the gut resistome (i.e., alpha diversity and abundance of clinically relevant ARGs) and microbiome composition (i.e., nonpareil sequence diversity and the relative abundances of the ESKAPEE pathogens *E. coli* and *K. pneumoniae*); (2) explore whether drinking water and sanitation conditions modify the relationship between animal exposure and the outcomes; and (3) examine the distribution of clinically relevant ARGs, their individual association with animal exposure, their genomic location (chromosomes or plasmids), and their linkage to ESKAPEE pathogens, with particular emphasis on beta-lactam resistance genes. This research was conducted as part of the ECoMiD (*Enteropatógenos, Crecimiento, Microbioma, y Diarrea*) study^41^ in Northwest Ecuador, in a region characterized by significant variability in WASH infrastructure and environmental exposures.^42^

## 2.0 Materials and methods

### 2.1 Ethics

The study protocol was approved by the institutional review boards of the University of Washington (UW; STUDY00014270), Emory University (IRB00101202), the University of California, San Francisco (21–33932), the Universidad San Francisco de Quito (USFQ; 2018– 022M), and the Ecuadorian Ministry of Health (MSPCURI000253–4). Written informed consent was obtained for all mothers both for their own participation, and on behalf of their children.

### 2.2 Sample and data collection

ECoMiD is a community-based birth cohort study in Ecuador designed to investigate the gut microbiome’s interactions with enteric infections and environmental conditions.^41^ Team members enrolled women in late pregnancy (∼37-weeks) and collected stool samples from mothers and their children starting at one-week post-birth and every three months thereafter, through 24 months of age. The ECoMiD study commenced enrollment in May 2019, with maternal samples collected within one week of consent. Following the initial enrollment phase, which continued through March 2020, the study experienced a pause due to the COVID-19 pandemic. Enrollment resumed in November 2020, and the final child samples were collected in December 2024. ECoMiD study design, sample collection and processing, and other details of data collection have been described previously.^41,42^ Briefly, stool samples were collected by caregivers in sterile containers. The samples were aliquoted by field staff and transported by in a portable liquid nitrogen tank to the Universidad San Francisco de Quito (USFQ), where they were stored at -80°C. DNA extraction was performed using the QIAamp Fast DNA Stool Mini Kit (Qiagen, Germantown, MD) with modifications described elsewhere.^42^ Frozen DNA samples were cold-shipped from USFQ to the University of Washington (UW) and Georgia Institute of Technology (Georgia Tech) and stored at -80°C until metagenomic sequencing.

Environmental and demographic variables were collected through structured questionnaires and fieldworker observations. The animal exposure score (calculated based on the FECEZ index^43^ - additional animal details in the SI), urbanicity classification, and socioeconomic status (SES) calculations are described in detail elsewhere.^42^ Briefly, we developed a score to measure exposure to animals and animal feces in all households enrolled in the ECoMiD study based on survey questions regarding animal ownership, animal-related behaviors, the presence of animal feces, and the presence of animals (score: 0-4).^42,43^ The score was categorized for the subset of samples in this study into none (0.00), low (0.00 - 0.55), medium (0.55 - 0.90), and high (>0.90) animal exposure, where the low, medium, and high cut-off values were based on tertiles calculated using the samples considered in this study. Communities were grouped into four categories with varying urbanicity: (1) Urban: the city of Esmeraldas (population ∼162,000), (2) Intermediate: the town of Borbón, (population ∼5,000), (3) Rural-road: rural villages that are accessible by road near Borbón, (populations ∼500-1000), and (4) Rural-river: rural villages that are mostly accessible by river with some limited car accessibility, (populations ∼200–700). An asset score was created as a proxy for SES using ownership data on 15 household possessions and multiple correspondence analysis. The asset score was divided into quartiles, representing SES, with 1 indicating the lowest and 4 the highest SES.^44^

Survey data on household environmental exposures were collected from mothers at ∼37-weeks of pregnancy and from children at 6- and 18-months of age. Consequently, animal exposure data for children were obtained from the nearest available timepoints: one-week data from the prenatal visit and three-month data from the 6-month visit. Drinking water, sanitation, and covariates in the linear models (e.g., community type, SES) were derived from survey responses at the pre-natal visit, as this information showed minimal variation over the course of the study. However, because piped water availability (days per week) was recorded for the week prior to each visit and this metric often varied over time, only samples from visits with this information (pre-natal visit, and 6- and 18-month visits) were included in the analysis.

### 2.3 Metagenomic sequencing and assembly

We performed shotgun metagenomic sequencing on 206 stool samples from 84 households enrolled in the ECoMiD study. Samples were collected from pregnant women at ∼37-week gestation (*n=53*) and their children at 1-week (*n=53*), 3-months (*n=20*), 6-months (*n=20*), and 18-months-old (*n=60*) (Table S1). A total of 53 samples from mothers and 93 samples from children (1-week, 3-months and 6-months) were sequenced at the Georgia Institute of Technology (Georgia Tech) Sequencing Core facility. These samples were selected based on collection prior to the COVID-19 pandemic project pause. At Georgia Tech, DNA extracts were subject to the Illumina DNA library preparation kit with 25-50ng of input and 5 PCR cycles followed by sequencing on an Illumina NovaSeq 6000 instrument using a 2 × 150 bp paired-end kit with 300 cycles. To expand our analysis of environmental exposures, we selected an additional 60 stool samples from 18-month-old children for metagenomic sequencing. Thirty of these samples were collected during the pre-COVID-19 enrollment period and corresponded to children who were already included in earlier time points (e.g., 1 week, 3 months, and 6 months). This allowed us to assess exposure over a longer developmental window (18 months). The remaining 30 samples were collected from children enrolled after the COVID-19 pause. These samples were selected to obtain a more balanced distribution of animal exposure across households. At UW, DNA extracts were prepared by the Microbial Interactions and Microbiome Center (mim_c) using the Illumina DNA library preparation kit with 10ng of input and 12 PCR cycles followed by sequencing on an Illumina NextSeq 2000. Three six-month-old samples were sequenced by both mim_c and Georgia Tech to ensure comparability of results (Figure S1). Raw data for the metagenomes was deposited in NCBI under BioProject PRJNA1225421.

The average shotgun metagenome read count was ∼23.8 (range 6-80) million reads per sample. Human reads were removed from the metagenomes using bbtagger v3.101^45^ while read trimming was performed using multitrim v1.2.6.^46^ For this, we first created a fasta file index for bmfilter and srprism using the commands *bmtool* and *srprism mkindex*, respectively. Then, we used blast v2.5.0^47^ to create a blast database using the command *makeblastdb* to remove the human reads with the command “bmtagger.sh”. Filtered metagenome reads were assigned taxonomy using kraken2 v2.1.3^48^ and diversity was calculated using the Nonpareil diversity index.^49,50^ Filtered reads were assembled using metaSpades v3.15.5^51^ with kmer sizes 21, 33, 55, 77, 99 and 127. The resulting assemblies were subject to gene calling using Prodigal v2.6.3^52^ and ARGs were annotated using the Comprehensive Antibiotic Resistance Database (CARD) v3.2.5^53,54^ using abricate v1.0.1.^55^ Contigs containing ARGs were taxonomically annotated with kraken2 v2.1.3 and these annotations were corroborated with blast. PlasX v0.0.0 was used for the detection and classification of plasmids within assemblies.^56^

### 2.4 Metagenome assembled genomes (MAGs) recovery, annotation, and dereplication

MAGs were recovered *de novo* from the assemblies of the first set of metagenomes sequenced at Georgia Tech (see section 2.2) after removing contigs less than 5000 bp using MetaBAT2 v2.12.1,^57^ and MaxBin2 v2.2.7^58^. MAGs were de-replicated using dRep v3.4.0^59^ at 95% ANI. MAG quality was evaluated with CheckM v1.2.2^60^ and taxonomy was determined with the Genome Taxonomy Database Toolkit (GTDB-Tk v2.3.2, database release 214).^61^ MAGs with a CheckM quality scores ≥ 50, calculated as “Quality = Completeness – (5 x Contamination)”, were selected, resulting in 285 MAGs used in further analyses. Prodigal v2.6.3^52^ was used to call genes and ARGs were annotated against the CARD database^53,54^ using ABRicate v1.0.1.^55^

### 2.5 Antimicrobial resistance genes (ARGs)

Trimmed metagenome reads were mapped to the CARD database v3.2.5 using the DIAMOND v2.0.15 aligner (--id 95 --query-cover 80 -e 0.00001 -k 1).^62^ We used the *BlastTab.seqdepth.pl* script from the enveomics toolkit to assess sequence depth for genes.^63^ Genome equivalent (GE) values were estimated using the MicrobeCensus tool v1.1.1.^64^ ARG relative abundance was estimated by normalizing the coverage of clinically relevant ARGs, as reported in Zhang et al., (2021)^65^, to genome equivalents (coverage/GE). In addition to the ARGs identified by Zhang et al., we expanded the list of clinically relevant ARGs to include all *bla*C_TX-M_, *bla*_SHV_, *bla*_OXA_, and *bla*_TEM_ alleles (see Table S2) due to the widespread prevalence and clinical significance of these β-lactamase genes.^66^ By encompassing all alleles of these genes, our analysis aims to capture the full spectrum of clinically relevant β-lactamase-mediated resistance, ensuring a comprehensive assessment of ARG diversity and abundance in our metagenomic samples. The alpha diversity of ARGs was defined as the number of unique clinically relevant ARGs (Table S2) per sample.

### 2.6 ESKAPEE pathogens

The presence and relative abundances of ESKAPEE pathogen strains were determined from the metagenomic data using the StrainGE toolkit v1.3.9^67^, which facilitates strain-level analysis and relative abundance quantification from metagenomes. First, a custom reference database was constructed using representative genome sequences of each of the ESKAPEE pathogens. All complete genome sequences for each ESKAPEE pathogen were downloaded from the NCBI RefSeq database^68^ to ensure comprehensive strain representation. All references were organized in a single directory using the script “prepare_strainge_db.py”. Then, reference sequences were kmerized using the *straingst kmerize* command. Redundant reference genomes were removed by computing the pairwise similarities between k-mer sets using the command *straingst kmersim* and references were clustered using *straingst cluster.* Because StrainGST compares the k-mer profiles of references in the database to the k-mers in the sample to identify close reference genomes to strains in a sample, we also kmerized the sample metagenome reads using the *straingst kmerize* command*. Straingst run* was used to list the identified reference strains and associated metrics. Concatenated reference FASTA files, containing a close reference genome for each strain identified in the sample, were created with *straingr prepare-ref*. Reference FASTA files for each strain identified in the sample were indexed with *bwa index*, and sequencing reads from each sample were aligned to the corresponding reference genome using *bwa mem*.^69^ Finally, we ran *straingr call* to call all strain variants in the samples and associated data, including strain name, relative abundance, and coverage in the metagenome sequencing data..

### 2.7 Statistical analyses

Statistical analysis was performed using R v4.4.1. We used multivariate linear regression models to determine associations between environmental exposures (animal exposures, sanitation facilities, drinking water access) and taxonomy diversity (Nonpareil sequence diversity), clinically relevant ARGs abundance, number of clinically relevant ARGs (ARGs diversity) and ESKAPEE pathogen relative abundance (i.e., *E. coli* and *K. pneumoniae*). Pair-wise significances were calculated with the Wilcoxon test.

#### 2.7.1 Exposure definitions

We considered the environmental exposures: (i) animal exposure score, (ii) combined sanitation & drinking water, and (iii) piped water availability for both mothers and children. Animal exposure score was defined using survey questions regarding animal ownership, animal-related behaviors, the presence of animal feces, and the presence of animals, as described above. For combined sanitation and drinking water, we divided the households into two categories: those with access to both contained sewage and piped water (CS&PW) versus “other”. Contained sewage was defined as households with sewer systems or septic tanks. Our definition of sanitation includes household services only, meaning that contained sewage did not indicate wastewater treatment. Piped water was defined as piped water reported as a household’s drinking water source. The “other” category included households who used latrines or other forms of sanitation, and bottled water, surface water, or rainwater for drinking. Categories were developed to reflect the variation in drinking water and sanitation conditions in our study area and that did not overlap with community type. Additionally, we evaluated associations with water availability, measured as the reported number of days households had piped water service during the week prior to sampling. Households were categorized as piped 7 days per week (d = 7), piped less than 7 days (0 < d < 7), and no piped water (d = 0).

#### 2.7.2 Outcome definitions

Outcome variables were: (i) clinically relevant ARG abundance, (ii) number of unique clinically relevant ARGs (ARG diversity), (iii) nonpareil sequence diversity, and (iv) the relative abundance of the ESKAPEE pathogens *E. coli* and *K. pneumoniae*. We created databases and ran strain-identifying analyses for all ESKAPEE pathogens but only *E. coli* (93%) and *K. pneumoniae* (52%) were detected in more than 20% of metagenomes. *A. baumannii* (3%) and *E. faecium* (11%) were also detected, but with insufficient prevalence. We therefore only included *E. coli* and *K. pneumoniae* in the linear model analyses.

#### 2.7.3 Multivariate linear regression models

Multivariate linear regression models were performed separately for mothers and children. We separated adults and children because the gut microbiome of children evolves rapidly during the first years of life, whereas adults typically have a more stable and diverse microbiome that is clearly distinguishable from children.^70^ In addition, children interact with their environments differently than mothers. For example, young children crawl, play on the ground, and put objects in their mouths. We used generalized linear models (GLM) with R’s glm() function for the maternal samples and controlled for SES and community type. Because child samples were longitudinal and involved repeated measures from the same individuals at different ages, we used generalized estimating equations (GEE) with the geeglm() function in R, accounting for within-child correlation.^44^ For the child models, we controlled for SES, community type, age, birth mode, and exclusive breastfeeding. Planned vaginal delivery was part of the inclusion criteria for the ECoMiD study; however, children delivered by unplanned emergency c-section were not excluded. Collinearity among predictors was evaluated by calculating variance inflation factors (VIF). Any covariates with a VIF greater than 4 were sequentially removed from the models to avoid multicollinearity. As a result, breastfeeding was removed since it was correlated with age.

#### 2.7.4 Interaction between WASH variables and animal exposure

We investigated whether drinking water and sanitation conditions modified the relationship between animal exposure and our outcomes of interest. Our effect modifier of interest was a binary variable indicating if the mother or child lived in a household with both contained sewage and piped water (CS&PW) versus a household with other combinations of water and sanitation systems (Other). We utilized the *waldtest* function in R to test for interaction between CS&PW and animal exposure. These interactions were assessed in adjusted models that included the covariates community type and SES for both mothers and children. Age and birth mode were also included in the child models.

## 3.0 Results

### 3.1 Overview of households included in this study and environmental exposures

Of the 84 households included in this analysis, 32 had piped water (38.1%) as their drinking water source. Other sources included bottled water, rainwater, surface water, protected wells, unprotected wells, and “other” sources. Most of the sanitation systems in the households were toilets that discharge into sewer systems (25.0%) or toilets that discharge to septic tanks (46.4%). Other sanitation systems included toilets that discharge to a pit latrine, improved and ventilated pit latrines, pit latrines with slab, pit latrines without slab and “other”. Among the 53 mothers and 84 children in our study, 39.6% of mothers and 28.6% of children resided in households equipped with both contained sewage systems and piped water (CS&PW). Regarding drinking water supply, 34.0% of mothers and 23.8% of children resided in households with piped water available 7 days a week, while 26.4% of mothers and 40.0% of children had no piped water. Among the mothers, 47.2% had zero animal exposure and 26.4% were classified as low, 18.9% as medium, and 7.5% as high animal exposure groups. Among the children, 37.9% had zero animal exposure and 20.9% were classified as low, 20.3% as medium, and 18.9% as high animal exposure (Table 1).

**Table 1:** Environmental exposure variables among mothers and children. The environmental variables in this study are animal exposure, drinking water and sanitation (DW&S), and water availability.

|  | Mothers | Children |
| --- | --- | --- |
| Households | 53 | 84 |
| Samples | 53 | 153 |
| **Sanitation** |  |  |
| Sewage containment<sup>1</sup> | 41/53 (77.4%) | 60/84 (71.4%) |
| No sewage containment | 10/53 (18.9%) | 19/84 (22.6%) |
| Not available (NA) | 2/53 (3.8%) | 5/84 (6.0%) |
| **Drinking Water** |  |  |
| Piped water | 27/53 (50.9%) | 32/84 (38.1%) |
| Not piped water | 26/53 (49.1%) | 52/84 (61.9%) |
| **DW&S** |  |  |
| Contained Sewage & Piped Water (CS&PW)<sup>2</sup> | 21/53 (39.6%) | 24/84 (28.6%) |
| Other | 30/53 (53.6%) | 58/84 (69.0%) |
| NA | 2/53 (3.8%) | 2/84 (2.4%) |
| **Animal exposure** |  |  |
| High | 4/53 (7.5%) | 29/153 (18.9%) |
| Medium | 10/53 (18.9%) | 31/153 (20.3%) |
| Low | 14/53 (26.4%) | 32/153 (20.9%) |
| Zero | 25/53 (47.2%) | 58/153 (37.9%) |
| NA | 0/53 (0.0%) | 3/153 (2.0%) |
| **Water availability<sup>3</sup>** |  |  |
| Not piped | 14/53 (26.4%) | 32/80 (40%) |
| Piped 0-6 days | 21/53 (39.6%) | 28/80 (35.0%) |
| Piped 7 days | 18/53 (34.0%) | 19/80 (23.7%) |
| NA | 0/53 (0.0%) | 1/80 (1.3%) |
| **Community type** |  |  |
| Urban | 14/53 (26.4%) | 16/84 (19.0%) |
| Intermediate | 21/53 (39.6%) | 29/84 (34.5%) |
| Rural-road | 15/53 (28.3%) | 32/84 (38.1%) |
| Rural-river | 3/53 (5.7%) | 7/84 (8.3%) |
| **Age (children)** |  |  |
| 1-week |  | 53/153 (34.6%) |
| 3-months |  | 20/153 (13.1%) |
| 6-monthsh |  | 20/153 (13.1%) |
| 18-months |  | 60/153 (39.2%) |

<sup>1</sup> Households with piped sewers or septic tanks
<sup>2</sup> Households with both piped sewers or septic tanks and piped drinking water
<sup>3</sup> Available only for samples at pre-natal visit, and 6- and 18-month visits

### 3.2 Number and abundance of clinically relevant ARGs

The number of clinically relevant ARGs in mothers (mean = 14, range = [2 - 29]), was significantly lower (p-value < 0.05) and less variable compared to children, (28.4 [7 - 95]). Clinically relevant ARG abundance ranged from 0.004 to 0.36 coverage/Genome Equivalents (GE) in mothers and from 0.007 to 3.13 coverage/GE in children. In mothers, the number of clinically relevant ARGs trended lower in both the low (β= -5.58 [95% CI: -11.46, 0.29]) and medium (β= -5.37 [-11.43, 0.68]) animal exposure tertiles compared to the high animal exposure group, although not statistically significant (Panel A of Figure 1 and Table S3). The number of clinically relevant ARGs in mothers with zero animal exposure was very similar to high animal exposure (β= -2.08 [-7.72, 3.56]). In children, both the number and abundance of clinically relevant ARGs increased when animal exposure decreased (Panels B and D of Figure 1 and Table S4). For example, the number of clinically relevant ARGs was 7.85 units higher in the low animal exposure group compared to the high animal exposure group (CI: -0.33, 16.03).

**Figure 1:**
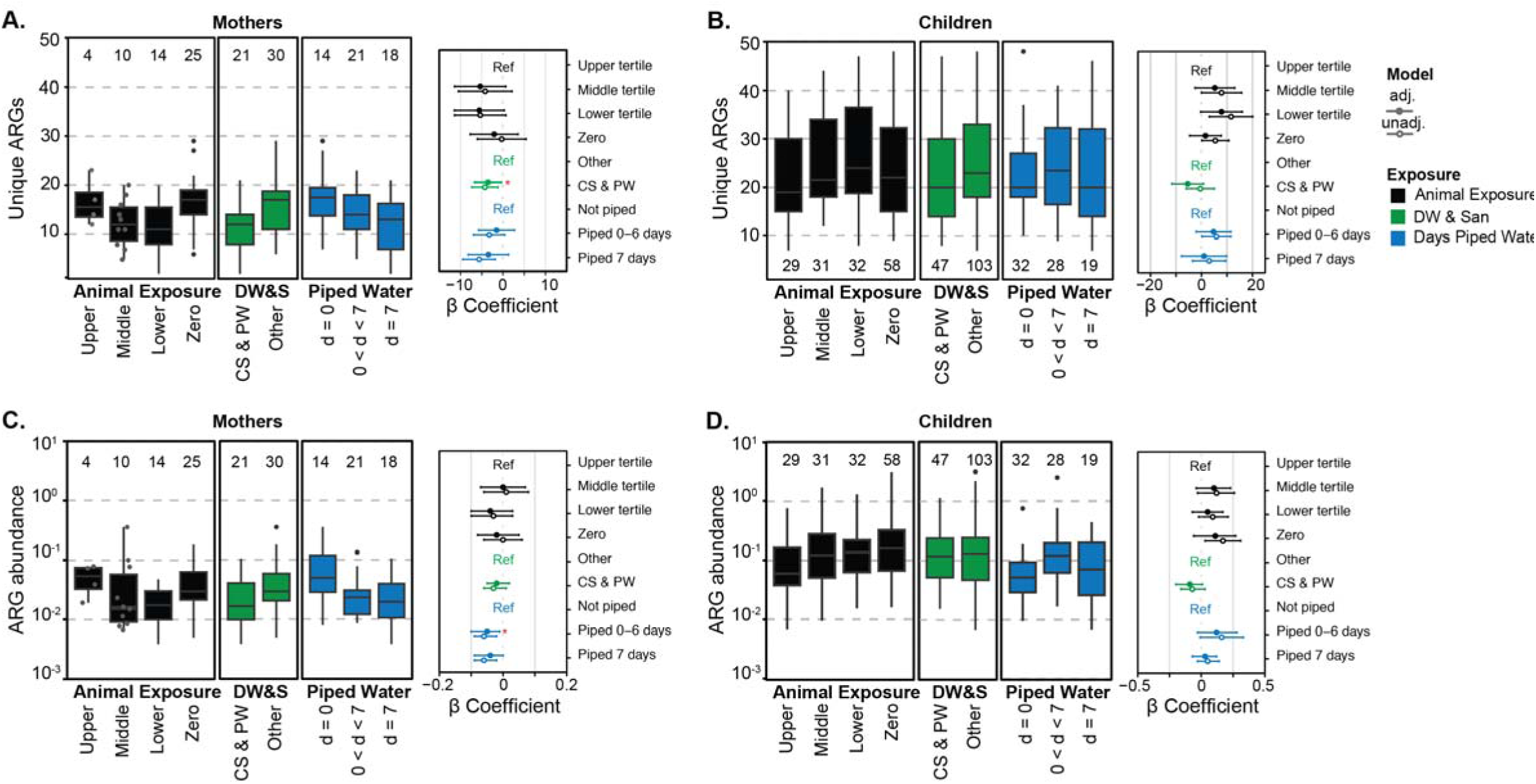
Antimicrobial resistance gene (ARG) diversity (number of unique clinically relevant ARGs) in mothers (A) and children (B) and normalized relative abundance of ARGs in mothers (C) and children (D), grouped by animal exposure levels (black), drinking water and sanitation (DW & San, green), and days of piped water availability (blue). The β coefficient for these variables (adjusted-close circle and unadjusted-open circle) are shown to the right of each panel with their 95% CI. Statistically significant β coefficients (p-value < 0.05) are marked with a red asterisk in adjusted models only. Animal exposure levels are zero exposure (zero), low exposure (lower tertile), medium exposure (middle tertile), and high exposure (upper tertile; reference in the linear model). Drinking water and sanitation (DW&S) categories were contained sewage and piped water (CS&PW) and other systems (other; reference in the linear model). Piped water availability categories are water available 7 days a week (d = 7), water available less than 7 days a week (0 < d < 7), and not piped systems (d = 0; reference in the linear model). Box plots show interquartile range (25th to 75th percentiles) with the median and whiskers extending to 1.5 times the interquartile range. Individual data points are shown in gray for n ≤ 10.

In mothers, the number of unique clinically relevant ARGs was significantly lower in households with CS&PW (β= -3.52 [-6.74, -0.30]) (Panel A of Figure 1). Households with greater access to piped water had lower clinically relevant ARG abundance compared to households without piped water (piped <7-day: -0.04 [-0.09, 0.00]). In children, both the number of ARGs (β= -5.42 [-11.64, 0.79]), and abundance of ARGS (β= -0.09 [-0.2, 0.01]) trended lower in households with CS&PW compared to households with other systems, though were not statistically significant.

### 3.3 Taxonomic diversity, E. coli and K. pneumoniae relative abundances

Sequence diversity (Nonpareil diversity) in mothers’ guts (mean = 18.4, range = [16.6-19.9]) was, on average, higher than for children (mean = 16.4, range = [14.5-18.5]) (Figure 2). Like the resistome, the diversity of child gut microbiomes spanned a wide range. Nonpareil diversity was lower in mothers with medium (β=-1.03 [-1.72, -0.35]), low (β= -1.29 [-1.96, - 0.63]), and no (β= -1.09 [-1.73, -0.45]) animal exposure compared to those with high exposure. There was no significant difference in sequence diversity when comparing mothers in households with CS&PW to those without (β= 0.02 [-0.39, 0.43]). In children, Nonpareil diversity was not associated with animal exposure and there were no significant differences between children in households with CS&PW and those without in the adjusted models (Animal exposure-middle tertile: β= 0.08 [-0.35, 0.52]; CS&PW: β= -0.1 [-0.37, 0.17]). Children with 7 days of piped water had significantly lower taxonomic diversity compared to no piped water (β=-0.64 [-1.14, -0.15]).

**Figure 2:**
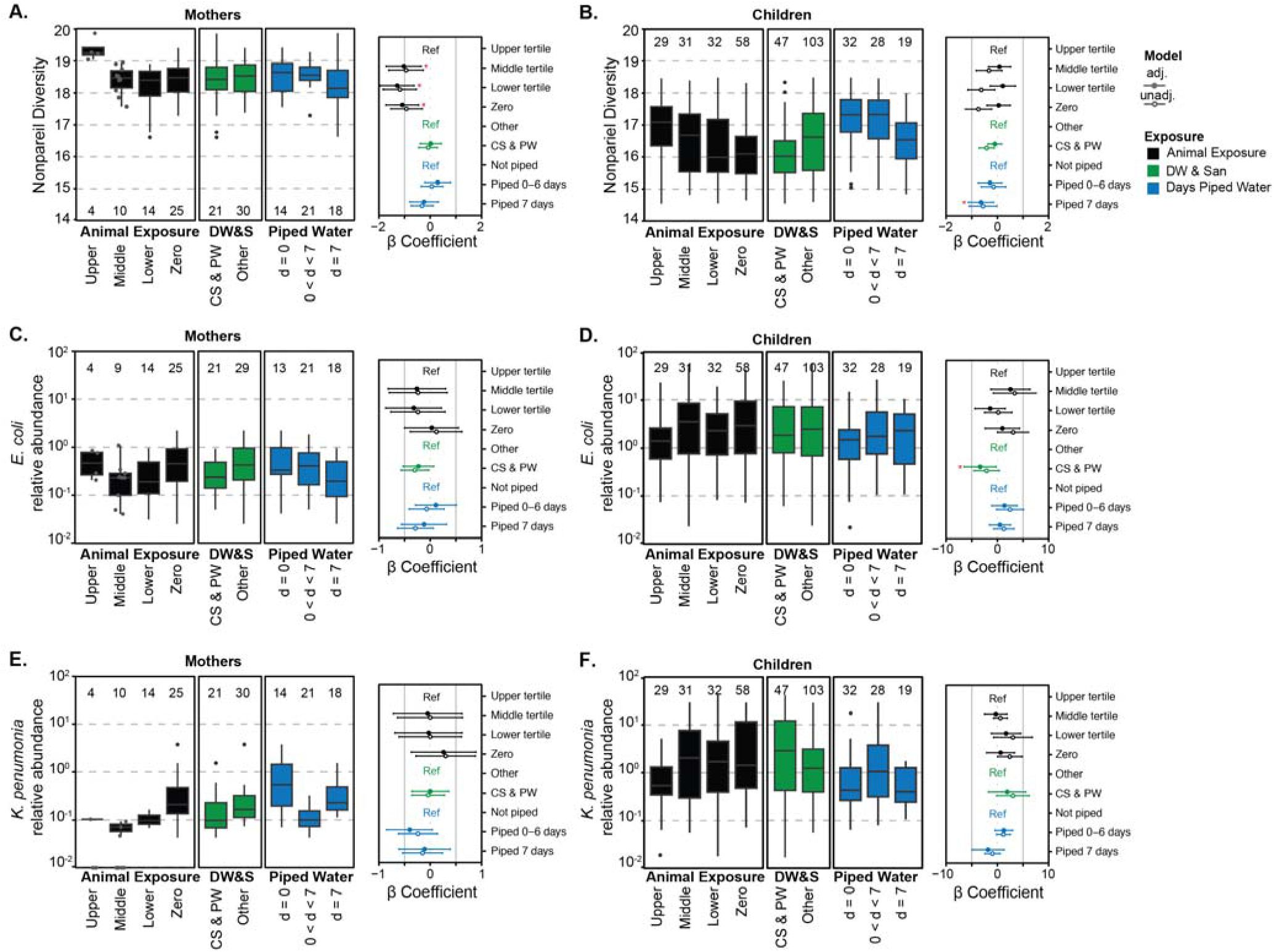
Nonpareil sequence diversity and relative abundance of *E. coli* and *K. pneumoniae* in mothers (A, C, E) and children (B, D, F), grouped by animal exposure levels (black), drinking water and sanitation (DW&S, green), and days of piped water availability (blue). The β coefficients for these variables (adjusted-close circle and unadjusted-open circle) are shown on the right side of each panel with their 95% CI. Statistically significant β coefficients (p-value < 0.05) are marked with a red asterisk for adjusted models only. Animal exposure levels are zero exposure (Zero), low exposure (Lower tertile), medium exposure (Middle tertile), and high exposure (Upper tertile; reference in the linear model). Drinking water and sanitation (DW&S) categories are contained sewage and piped water (CS&PW) and other systems (Others; reference in the linear model). Piped water availability categories are water available 7 days a week (d = 7), water available less than 7 days (0 < d < 7), and not piped systems (d = 0; reference in the linear model). One mother sample was removed for relative abundance of *E. coli* since it was greater than two standard deviations away from the mean. Box plots show interquartile range (25th to 75th percentiles) with the median and whiskers extending to 1.5 times the interquartile range. Individual data points are shown in gray for n ≤ 10.

The relative abundances of *E. coli* and *K. pneumoniae* in mothers (*EC*: mean: 0.5% (range: 0-19.8%); *KP:* 0.2 (0-3.8) %) were lower than in children (*EC*: 5.4 (0-53.4) %; *KP*: 2.9 (0-43.6) %). *E. coli* relative abundance in the maternal samples with zero animal exposure was not different from those with high animal exposure (β= 0.03 [-0.5, 0.55]). Similar to ARGs in children, *E. coli* abundance was higher in the zero-animal exposure compared to higher animal exposure group in the unadjusted model (β= 3.06 [-0.01, 6.13]) although not significant in the adjusted model (β= 0.97 [-2.39, 4.32]). *E. coli* abundance in children from households with CS&PW was lower than those with other systems (β= -3.37 [-6.49, -0.24]). *K. pneumoniae* relative abundance was highest in mothers with zero animal exposure compared to other exposure groups and not statistically different from mothers with high animal exposure (β= 0.26 [-0.37, 0.89]). *K. pneumoniae* relative abundance was higher in children compared to mothers and showed no consistent trends across exposure categories.

### 3.4 Differential associations between animal exposure by WASH access

The associations between animal exposure and the microbial outcomes in mothers follow similar trends between households with and without CS&PW (Panel A of Figure 3). However, some correlations were stronger than others. For example, the number of clinically relevant ARGs was lower in households with low and medium animal exposure, compared to households with high animal exposure, in both groups (households with and without CS&PW) (Table S5). However, associations were significant only in households without CS&PW (e.g., no CS&PW/animal expo-low tertile: β= -10.31 [-19.29, -1.34]; CS&PW/animal expo-low tertile: β=-5.42 [-13.56, 2.73]). The number of clinically relevant ARGs, sequence diversity, and relative abundance of *E. coli* and *K. pneumoniae* were higher in children with less animal exposure in households with CS&PW but not in households without CS&PW (i.e., “Other” systems) (Table S6).

**Figure 3:**
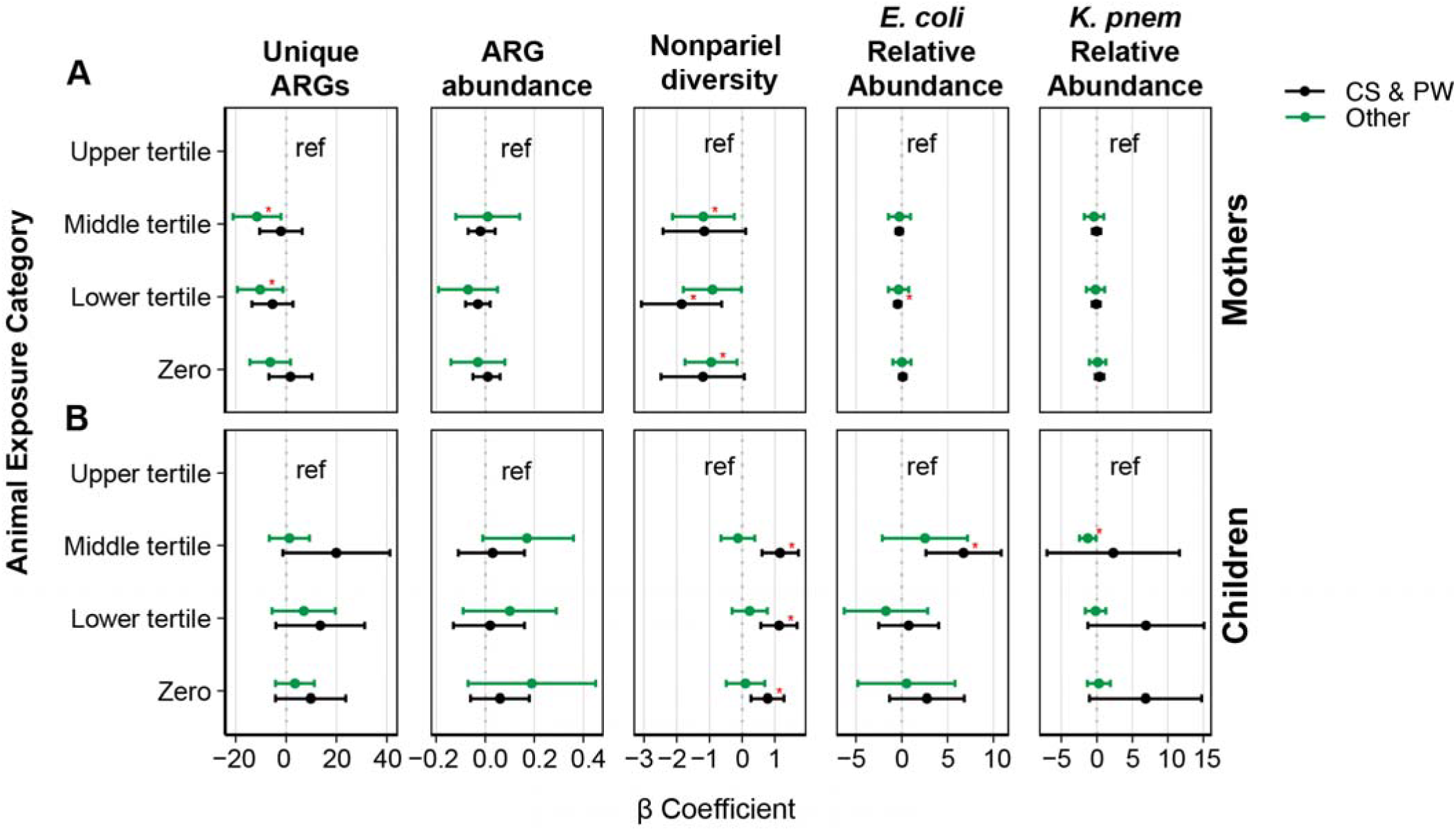
Adjusted animal exposure β coefficients stratified by drinking water and sanitation (DW&S) for mothers (A) and children (B). Each plot presents β coefficients (with 95% confidence intervals) between animal exposure categories (Upper tertile, Middle tertile, Lower tertile, and Zero exposure) and the outcomes: Number of clinically relevant ARGs (Unique ARGs), ARGs abundance (coverage/GE), nonpareil sequence diversity, and the relative abundance (%) of *E. coli* and *K. pneumoniae* in households with contained sewage and piped water (CS&PW-black) and without CS&PW (Other-green). Statistically significant β coefficients (p-value < 0.05) are marked with a red asterisk.

### 3.5 Distribution of clinically relevant ARGs and their ESKAPEE pathogen hosts

Genes that indicate resistance to beta-lactams, specifically *bla*_CTX-M_, *bla*_OXA_, *bla*_SHV_, and *bla_TEM_*, were found in contigs from all age groups: 37 weeks of pregnancy (*n*=6/53), one-week-old (*n=*21/53), three-months-old (*n=*6/20), six-months-old (*n=*5/20), and 18-months-old (*n=*29/60). ESBL genes were consistently found in mothers and children across age groups although the majority of ARG containing contigs were identified in children (Figure 4A). There was no association between clinically relevant ARG-containing contigs and animal exposure (low exposure tertile in mothers: β= -0.65 [-1.94, 0.66], p-value = 0.31; low exposure tertile in children: β= 1.29 [-0.24, 1.82], p-value = 0.10). Similarly, the ESKAPEE MAGs were found across age groups and were not associated with animal exposure status (low exposure tertile in mothers: β= 0 [-0.006, 0.007], p-value = 0.88; low exposure tertile in children: β= 0 [-0.03, 0.03], p-value = 0.92). *E. coli* and *K. pneumoniae* had the highest coverage and were present in most samples whereas the coverage of *A. baumannii* was zero in all mother samples (Figure 4 C). In mothers and children, the ESKAPEE pathogen MAGs harbored diverse clinically relevant ARGs conferring resistance to multiple antibiotic classes. Key beta-lactam resistance genes include *bla*_CTX-M-15_, *bla*_SHV-106_, *bla*_TEM-1_, and *bla*_OXA-120_, which confer resistance to penicillins, cephalosporins, and carbapenems, and were found in the *K. pneumoniae*, and *A. baumannii* MAGs. Aminoglycoside resistance genes, such as *aac*(*6’)-I* and *aph(6)-I*, confer resistance to drugs like gentamicin and kanamycin and were found in *K. pneumoniae* and *E. faecium*. A gene for fluoroquinolone resistance (*qnrB1*) was found in *K. pneumoniae*, while a gene conferring macrolide resistance (*ermB*) was present in the *E. coli* MAG.

**Figure 4:**
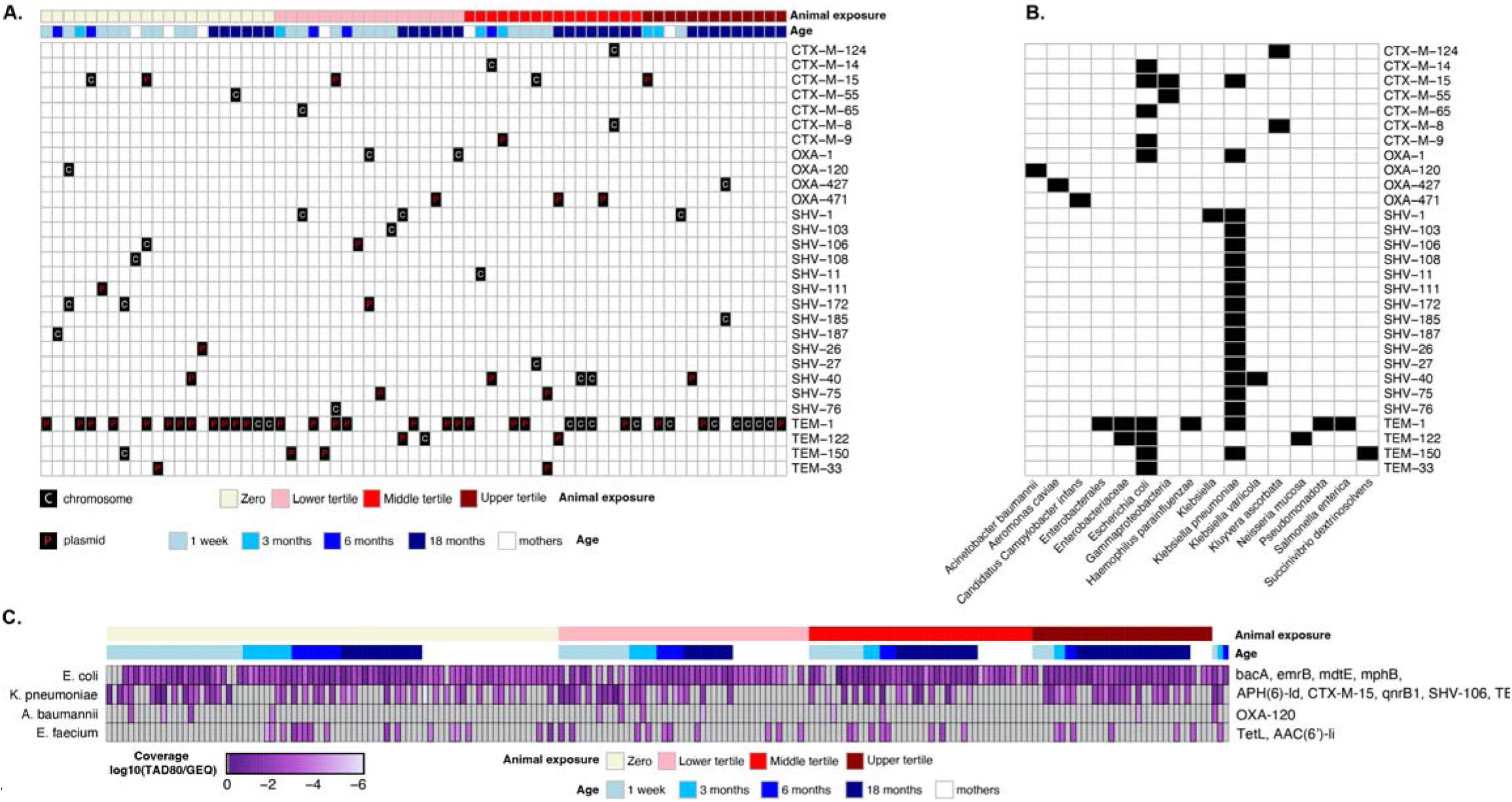
(A) Heatmap illustrating the presence of beta-lactamase genes (*bla*_CTX-M_, *bla*_OXA_, *bla*_SHV_, and *bla_TEM_*, families) on contigs assembled from metagenome sequences across samples. Rows represent specific antimicrobial resistance genes (ARGs) assembled in contigs, and columns represent individual samples, categorized by age [mothers (white) and children (shades of blue) at 1 week, 3 months, 6 months, and 18 months] and organized by animal exposure levels [Zero (light yellow), Lower Tertile (pink), Middle Tertile (red), and Upper Tertile (dark red)]. ARGs are classified as chromosomal (“C”) or plasmid-borne (“P”). (B) Taxonomic classifications of contigs carrying beta-lactamase genes ( *bla*_CTX-M_, *bla*_OXA_, *bla*_SHV_, and *bla_TEM_*, families) across samples. Rows correspond to specific ARGs, and columns indicate bacterial taxa identified at the highest resolution by Kraken2. Black squares indicate the presence of ARGs within each taxon. (C) Heatmap of MAGs identified as ESKAPEE pathogens containing clinically relevant ARGs. Abundances (log10-transformed coverage) are shown across samples stratified by age and organized by level of animal exposure. Rows represent the MAG (*E. coli, K. pneumoniae, A. baumannii, and E. faecium*), and columns represent individual samples.

Plasmid-borne ARGs, which are easily transmitted via horizontal gene transfer, were more common than chromosomal ARGs in our dataset; we found 48 ARGs on plasmids and 40 on chromosomes (Figure 4A). This was particularly true for the genes *bla*_CTX-M-15_, *bla*_OXA-471_, and *bl*a_SHV-40_. In contrast, ARGs such as *bla*_SHV-1_ and *bla*_OXA-1_ were primarily located on chromosomes. In general, the *bla*_CTX-M_, *bla*_OXA_, *bla*_SHV_, and *bla_TEM_* genes are widely distributed across both chromosomal and plasmid DNAs. Consistent with the association of ESKAPEE pathogen MAGs with beta-lactam resistance, *bla*_CTX-M_ and *bla*_TEM_ genes were mainly identified in *E. coli* and *K. pneumoniae*, while *bla*_OXA_ and *bla*_SHV_ were typically found in *K. pneumoniae*. Only a few *bla*_CTX-M_, *bla*_OXA_, and *bla_TEM_* genes were detected in other bacteria, including *Candidatus* Campylobacter infans and *Salmonella enterica* (Figure 4B).

## 4.0 Discussion

We found evidence that exposure to animals may be associated with increased number of clinically relevant ARGs and taxonomic diversity in adults (mothers), but not in children. Our findings align with existing research indicating that interactions at the human-animal interface can significantly influence the human microbiome and resistome.^21,23,71^ We did not find differences in the relative abundance of *E. coli* or *K. pneumoniae* between mothers with low and high animal exposure, and mothers with zero animal exposure trended towards higher relative abundances of *K. pneumoniae* compared to exposed mothers. This aligns with evidence that, although *K. pneumoniae* is common in both humans and animals, direct zoonotic transmission appears likely limited.^72^ Our findings differ from studies linking animal contact to increased *E. coli* colonization in humans^73^ and from those showing identical *E. coli* strains in pets and owners.^74^ This discrepancy may be due to our relatively small high-exposure group, which limited our ability to detect subtle changes in *E. coli* abundance.

Previous studies have demonstrated that livestock^10^ and pets^11,12^ harbor a wide array of ARGs, which can be transferred to humans through food, direct contact with feces, or environmental pathways.^75,76^ Environmental dissemination can occur when bacteria and antibiotic residues from animal production spread through manure, impacting environmental bacterial populations.^76^ Consequently, the environment can become reservoirs of resistance, further facilitating the reintroduction of antimicrobial resistance into human and animal reservoirs.^77,78^ The lack of difference between “zero” and high animal exposure levels in mothers in this study suggests that indirect exposure routes (e.g., from environmental fecal contamination) may play an important role in disseminating ARGs. Defining "zero exposure" is particularly challenging in settings where animals are prevalent within the community, as environmental reservoirs (e.g., soil and water bodies) can serve as pathways for ARG transmission via exposure to animal waste, independently of direct animal contact.^79^ This complicates the task of accurately defining exposure levels.^80^

Interestingly, we did not observe similar associations between animal exposures and ARG diversity and abundance or ESKAPEE pathogen carriage in children. The child resistome and microbiome were more variable compared with mothers. The abundance of ARGs, *E. coli*, and *K. pneumoniae* trended higher in the zero and lower animal exposure groups compared to the high animal exposure group, though not statistically significant. Although the challenges of defining "zero exposure" could again explain why these abundances are higher at zero animal exposure; the dynamic nature of the gut microbiome during infancy may also contribute.^81,82^ Early-life factors such as breastfeeding and dietary transitions significantly influence microbial colonization. For example, breastfeeding promotes the growth of beneficial bacteria like *Bifidobacterium,*^83^ while introducing solid foods leads to changes in the composition of the gut microbiome.^83,84^ The rapidly changing microbial community in children compared to adults may explain the lack of observed associations between animal exposure and the microbial outcomes in the infant population.

Improved sanitation and water infrastructure (i.e., CS&PW) trended towards lower diversity and abundance of clinically relevant ARGs and a lower relative abundance of *E. coli* in mothers and children, highlighting the role that water and sanitation services play in mitigating microbial transmission and reducing the prevalence of ARGs in the gut. These findings are consistent with previous research showing that access to improved water and sanitation correlated with lower ARG abundance in human gut metagenomes aggregated from 26 countries.^85^ Improved WASH conditions can also diminish the incidence of infections, thereby reducing the necessity for antibiotics and potentially limiting the spread of ARGs. In a randomized controlled trial of WASH interventions in Kenya and Bangladesh, WASH interventions were associated with 10-14% lower antibiotic consumption in children in Bangladesh.^86^ Improving water and sanitation facilities could decrease the prevalence of ARGs in the environment, reducing human and animal exposure.

Mothers in households with more days of available piped water had lower clinically relevant ARG abundance compared to households without piped water. In regions where piped water supply is unreliable, residents often resort to alternative water sources, such as untreated surface water or storage systems, to meet their daily needs.^87,88^ Secondary storage containers typically reduce drinking water quality through recontamination.^89^ In addition, intermittent water supply (IWS), characterized by the delivery of water for limited hours or days, can significantly impact water quality through contamination in the pipelines.^90,91^ IWS allows for stagnation and depressurization, creating conditions favorable for biofilm growth on pipe surfaces as well as intrusion of environmental bacteria.^92^ While studies specifically investigating the impact of IWS on the microbial communities in the human gut microbial are limited, existing research indicates that chlorination to improve drinking water quality has minor impacts on the resistome and microbiome.^93^

While previous research indicates that improved WASH infrastructure can reduce microbial transmission and the spread of ARGs,^85^ we did not find evidence that adequate drinking water and sanitation infrastructure mitigate the impact of animal exposure on the maternal gut microbiome. Conversely, in households with CS&PW systems, children with lower animal exposure had higher number of clinically relevant ARGs and taxonomic diversity as well as higher relative abundance of *E. coli* and *K. pneumoniae*. This counterintuitive finding may indicate that there are alternative environmental contamination pathways contributing to ARG occurrence in children besides WASH and animal exposure. Beyond improving WASH infrastructure, additional measures are likely necessary to address environmental exposures affecting children’s gut microbiomes. This may include interventions targeting behavioral practices and broader community-level factors to reduce the burden of antimicrobial resistance.^94^ Our findings in the stratified animal exposure analysis may be partially attributed to the limited number of samples with high animal exposure in these settings, which affects the statistical power of our analysis. Therefore, the observed patterns should be interpreted cautiously, and further studies with larger sample sizes are necessary to validate these findings.

The detection of clinically relevant ARGs across all age groups highlights the widespread nature of beta-lactamase genes such as *bla*_CTX-M_, *bla*_OXA_, *bla*_SHV_, and *bla_TEM_*. This finding aligns with previous research indicating that certain beta-lactamase genes are prevalent across populations.^95^ The higher prevalence of ARGs in younger children compared to mothers aligns with existing research,^96,97^ even when infants have not been exposed to antibiotics.^98^ The high levels of *Gammaproteobacteria*, which are common early gut colonizers and often carry resistance genes, may explain this increased ARG load in the child gut, though the exact source of these resistant strains (whether from the environment, other individuals, or the mother) remains unclear.^98^

The greater proportion of plasmid-borne compared to chromosomal ARGs underscores the critical role of HGT in facilitating the rapid dissemination of resistance traits between bacterial populations. Plasmids, as mobile genetic elements, can transfer ARGs between different bacterial species, increasing the spread of antimicrobial resistance.^99^ This interspecies transfer is particularly concerning in clinical settings, where it can lead to the emergence of pathogens with resistance to multiple antibiotics. For instance, plasmids have been identified as key vectors in the spread of ESBL genes among *Enterobacteriaceae*, leading to infections that are difficult to treat.^100^ In this study, *bla*_CTX-M-15_, *bl*a_OXA-471_, and *bla*_SHV-40_ genes were predominantly plasmid-borne, while ARGs like *bla*_SHV-1_ and *bla*_OXA-1_ were primarily chromosomal. Understanding the distinct roles of plasmid-borne and chromosomal ARGs in the propagation of antibiotic resistance is crucial for developing targeted strategies to combat the rise of multidrug-resistant bacteria.^101^

*E. coli* and *K. pneumoniae* were the most frequently identified ESKAPEE pathogens in the metagenomes, with MAGs and StrainGE-identified strains detected in most samples. In contrast, *A. baumannii* MAGs were notably absent in all mother samples. The difference in relative abundance of ESKAPEE pathogens may reflect differences in environmental reservoirs or host susceptibility among age groups. *E. coli* is a common inhabitant of the human gut microbiota.^102^ Similarly, *K. pneumoniae* is known to colonize the gastrointestinal tract, with prevalence rates varying geographically.^103^ In contrast, *A. baumannii* is primarily recognized as an opportunistic pathogen associated with healthcare settings.^104^ Among the ESKAPEE MAGs, *bla*_CTX-M_ and *bla*_TEM_ genes were mainly identified in *E. coli* and *K. pneumoniae*, while *bla*_OXA_ and *bla*_SHV_ were typically found in *K. pneumoniae*. This observation aligns with previous reports highlighting the association of these genes with ESBL production in these pathogens.^105^ Detecting these genes in pathogens of significant clinical concern highlights the importance of continuous surveillance and targeted interventions to mitigate the spread of resistance.

Applying StrainGE for targeted strain detection and assembling MAGs for ESKAPEE pathogens allowed us to directly compare these approaches. StrainGE is a metagenomic tool built to identify and characterize low-abundance strains within complex microbial communities using short-read metagenomic datasets. It has primarily been used for identifying *E. coli* strains.^67^ In our study, StrainGE was specifically used to identify and quantify the relative abundance of other ESKAPEE pathogens in addition to *E. coli*. The relative abundance and presence of ESKAPEE pathogens as well as overall trends in animal exposure were consistent between StrainGE and assembled MAGs. Both methods consistently detected the presence of *E. coli*, *K. pneumoniae*, *E. faecium*, and *A. baumannii* across samples, with *E. coli* and *K. pneumoniae* present in more samples and at greater relative abundances. These findings suggest that despite methodological differences, StrainGE and MAG-based approaches yield comparable insights into the prevalence and dominance of clinically relevant pathogens in the gut microbiome. The consistent results obtained from these two approaches supports StrainGE as a powerful tool for characterizing other bacterial species beyond *E. coli*. We note however that the effectiveness of StrainGE is closely linked to the quality and comprehensiveness of its reference genome database.

We applied a well-considered animal exposure assessment by calculating a score using the framework developed for the FECEZ index.^43^ However, accurately categorizing animal exposure levels can be particularly challenging in communities where animals are common, and exposure can occur in multiple settings (additional animal details in the supporting information). This may result in misclassification, which could impact the associations between animal exposure and microbiome or resistome profiles. In our study, the animal exposure score encompassed a range of animals commonly found in Ecuadorian households, including companion animals such as dogs and cats, as well as livestock like poultry, pigs, and cows. Companion animals and livestock differ in their antibiotic exposure and potential to contribute to AMR transmission. Pets often receive antibiotics for medical treatments, and close contact with humans (e.g., petting, licking, and sharing living spaces) facilitates the bidirectional transmission of resistant bacteria.^106^ In contrast, livestock are frequently administered antibiotics not only for therapeutic purposes but also for growth promotion and disease prevention.^107^ This widespread use contributes to the selection of resistant bacteria within these animal populations, which can be transmitted to humans through direct contact, consumption of contaminated products, or environmental pathways. The inclusion of both companion animals and livestock in our exposure assessment reflects the diverse animal-human interactions present in the study communities. However, the varying degrees of antibiotic exposure and human contact among these animal types underscore the complexity of AMR transmission dynamics. Additionally, the lack of direct sampling from animals prevents a comprehensive investigation of transmission pathways between animals and humans. While evidence suggests resistant bacteria transmission occurs between household animals and humans in this region,^108^ we were not able to investigate direct AMR transmission without animal microbiome and resistome data. Future studies incorporating direct sampling of both pets and livestock, along with detailed antibiotic usage data, would enhance our understanding of their respective roles in the dissemination of antimicrobial resistance. Finally, the relatively small sample size, particularly in the high animal exposure groups, limits the statistical power of our study.

## 5.0 Acknowledgments

We thank the participants of the ECoMiD study and the dedicated field staff who administered the surveys and collected samples. Sequencing support for 60 samples was provided by the University of Washington Microbial Interactions & Microbiome Center (mim_c). I.C. was supported under the ASEE e-fellows program Federal Award 2127509. E.R.F was supported by the University of Washington Interdisciplinary Center for Exposures, Diseases, Genomics & Environment under grant P30 ES007033. This work was funded by the National Institutes of Health (R01AI137679 and R01AI162867). The content is solely the responsibility of the authors and does not necessarily represent the official views of the National Institutes of Health.

## 6.0 Data Availability

All fastq files generated in this study have been deposited in the Sequence Read Archives under BioProject PRJNA1225421.

