## Supporting Information for "Environmental Exposures and the Human Gut Resistome in Northwest Ecuador"

**ECoMiD Authorship Group:**

Principal Investigators/Co-Investigators

Karen Levy^1^, Joseph N.S. Eisenberg^2^, Gwenyth O. Lee^3^, Gabriel Trueba^4^, Benjamin F. Arnold^5^, Konstantinos T. Konstantinidis^6^, William Cevallos^7^

Field data collection subgroup

Adriana Lupero^4^, Mauricio Ayoví^4^, Molly K. Miller-Petrie^1^

Data management subgroup

Jesse Contreras^2^, Jessica Uruchima^2^

Laboratory Analysis subgroup

- *Lab coordination*: Christine Fagnant-Sperati^1^, Gabriela Vasco^8^, Stuart Torres^4^
- *Gut microbiome*: Janet Hatt^6^, Ana Duran-Viseras^6^, Kelsey Jesser^1^

Animal Exposure Subgroup

- *Qualitative & Survey Data*: April Ballard^10^, Bethany Caruso^11^, Betty Corozo^12^

^1^ University of Washington, Department of Environmental and Occupational Health Sciences, Seattle, WA, USA

^2^ University of Michigan, Department of Epidemiology, Ann Arbor, MI, USA

^3^ Rutgers University, Global Health Institute and Department of Biostatistics and Epidemiology, New Brunswick, New Jersey, USA

^4^ Universidad San Francisco de Quito, Instituto de Microbiologia, Quito, Ecuador

^5^ University of California San Francisco, Proctor Foundation and Department of Opthamology, San Francisco, CA, USA

^6^ Georgia Institute of Technology, School of Civil & Environmental Engineering, Atlanta, GA, USA

^7^ Universidad Central del Ecuador, Instituto de Biomedicina, Quito, Ecuador

^8^ Universidad Central del Ecuador, Facultad de Ciencias Médicas, Carrera de Medicina, Quito, Ecuador

^9^ Universidad de las Americas, Facultad de Medicina, Carrera de Medicina, Quito, Ecuador

^10^ Georgia State University, Department of Population Health Sciences, Atlanta, GA, USA

^11^ Emory University, Department of Global Health, Atlanta, Georgia, USA

^12^ Universidad Técnica Luis Vargas Torres de Esmeraldas, Esmeraldas, Ecuador

^13^ South Florida University, Department of Integrative Biology, Tampa, FL, USA

^14^ Universidad San Francisco de Quito, Escuela de Salud Pública y Nutrición

**Household Animal Presence and Environmental Fecal Contamination**

**In the study communities along Ecuador’s northwestern coast, a wide array of domestic and semi-domestic animals is common in and around household compounds. Households frequently keep poultry (primarily chickens) that are allowed to roam freely throughout yards and communal pathways, depositing feces wherever they forage^1^ with 22 of 84 households owning chickens in this study**. Dogs and cats, whether owned (46 of 84 households and 20 of 84 households, respectively) or free-roaming strays, move in and out of houses and play areas, further spreading animal feces across domestic spaces. Livestock such as pigs are often penned near the home but sometimes released during the day to scavenge (7 households own in this study), while larger stock, including cattle and horses, graze in nearby fields that children traverse daily (1 household own in this study). Animal feces management practices reported by mothers typically involve rinsing or sweeping droppings into open drainage ditches or into adjacent yards rather than **containing or treating them, leading to persistent environmental contamination around homes and communal areas. Even households without animal ownership observed feces tracking into living spaces because roaming animals defecate indiscriminately, highlighting that exposure pathways extend beyond owned-animal contacts to a broader community-level norm of free-range animal husbandry. Additional details on animal exposure in this region are available elsewhere.^1–3^**

**Metagenomic Results Considering Sequencing Coverage**

To ensure our results are robust and not due to differential coverage (or differences in the fraction of diversity that was sequenced), we employed the Nonpareil tool to assess and standardize coverage levels. Nonpareil estimates the redundancy and diversity of metagenomic datasets without reliance on reference databases, providing a robust measure of sequencing coverage.^4,5^ We calculated the Nonpareil coverage (Npc) for each metagenome. The Npc values for maternal samples ranged from 0.68 to 0.94, while those for child samples ranged from 0.67 to 0.99. To ensure comparability, we excluded samples where the Npc difference exceeded 0.2 units within each group, based on the metagenome with the highest Npc, resulting in the removal of five maternal and four child samples. We analyzed the impact of exposures in the reduced dataset using the same regression models described in the main text. The results were consistent with our original findings, indicating that sequencing coverage differences did not influence our results. As an additional check, we applied a coverage standardization script based on Nonpareil estimates^6^ to normalize ARG abundance data. This approach adjusts for sequencing depth variations, ensuring that observed differences in ARG profiles are not artifacts of sequencing effort. Post-standardization, the ARG outcomes were nearly identical to the initial results, reinforcing that our findings are not due to differences in the diversity that was sequenced.

| **Table S1: Samples included in this study.** | | | | | |
| --- | --- | --- | --- | --- | --- |
| **Household ID** | **Age** | | | | |
|  | **37w** | **01w** | **03m** | **06m** | **18m** |
| **1001** |  | * | * | * | * |
| **1002** | * | * | * | * | * |
| **1003** | * | * | * | * | * |
| **1004** | * | * | * | * | * |
| **1101** | * | * | * | * | * |
| **1102** | * | * | * | * | * |
| **1103** | * | * | * | * | * |
| **1104** | * | * | * | * | * |
| **1106** | * | * | * | * | * |
| **2002** | * | * | * | * | * |
| **2003** | * | * | * | * | * |
| **2202** | * |  | * | * |  |
| **2204** | * | * | * | * | * |
| **2301** | * | * | * | * | * |
| **2302** | * | * | * | * | * |
| **3001** | * | * | * | * | * |
| **3101** | * | * | * | * | * |
| **3201** | * | * | * | * | * |
| **7001** | * | * | * | * | * |
| **7001** | * | * | * | * |  |
| **1007** | * | * |  |  | * |
| **1107** | * | * |  |  | * |
| **2009** | * | * |  |  | * |
| **2101** | * | * |  |  | * |
| **2205** | * | * |  |  | * |
| **2206** | * | * |  |  | * |
| **2306** | * | * |  |  | * |
| **3004** | * | * |  |  | * |
| **3006** | * | * |  |  | * |
| **3102** | * | * |  |  | * |
| **6001** | * | * |  |  | * |
| **6006** | * | * |  |  | * |
| **8001** | * | * |  |  |  |
| **8002** | * | * |  |  |  |
| **8003** | * | * |  |  |  |
| **2001** | * | * |  |  |  |
| **2004** | * | * |  |  |  |
| **2008** | * | * |  |  |  |
| **2102** | * | * |  |  |  |
| **2103** | * | * |  |  |  |
| **2201** | * | * |  |  |  |
| **2304** | * | * |  |  |  |
| **2305** | * | * |  |  |  |
| **2308** | * | * |  |  |  |
| **2311** | * | * |  |  |  |
| **4002** | * | * |  |  |  |
| **1005** | * | * |  |  |  |
| **1006** | * | * |  |  |  |
| **1011** | * | * |  |  |  |
| **1012** | * | * |  |  |  |
| **3005** | * | * |  |  |  |
| **3103** | * | * |  |  |  |
| **6002** | * | * |  |  |  |
| **6004** | * | * |  |  |  |
| **2213** |  |  |  |  | * |
| **3007** |  |  |  |  | * |
| **3015** |  |  |  |  | * |
| **3108** |  |  |  |  | * |
| **3110** |  |  |  |  | * |
| **3112** |  |  |  |  | * |
| **3115** |  |  |  |  | * |
| **3012** |  |  |  |  | * |
| **3107** |  |  |  |  | * |
| **9009** |  |  |  |  | * |
| **5001** |  |  |  |  | * |
| **9006** |  |  |  |  | * |
| **2317** |  |  |  |  | * |
| **9005** |  |  |  |  | * |
| **1116** |  |  |  |  | * |
| **2119** |  |  |  |  | * |
| **7003** |  |  |  |  | * |
| **2219** |  |  |  |  | * |
| **3002** |  |  |  |  | * |
| **2320** |  |  |  |  | * |
| **3011** |  |  |  |  | * |
| **9003** |  |  |  |  | * |
| **9001** |  |  |  |  | * |
| **2012** |  |  |  |  | * |
| **2322** |  |  |  |  | * |
| **4006** |  |  |  |  | * |
| **2312** |  |  |  |  | * |
| **6009** |  |  |  |  | * |
| **9002** |  |  |  |  | * |
| **7011** |  |  |  |  | * |

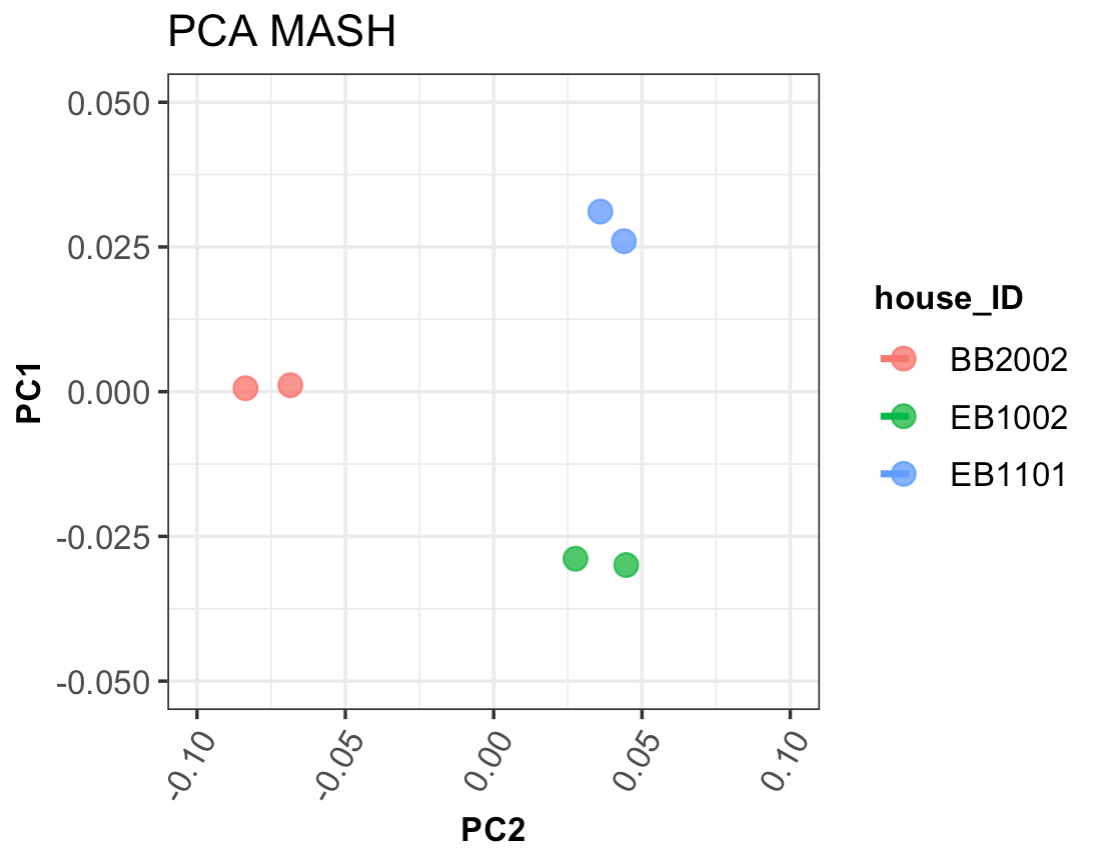
**Figure S1:** PCA of MASH distance of three 6-month samples sequenced at both Georgia Tech and the University of Washington.

| **Table S3:** Linear model results for the microbial outcomes with animal exposure, DW & San, and piped water availability in mothers. | | | | | | |
| --- | --- | --- | --- | --- | --- | --- |
|  |  | **Unadjusted** | | **Adjusted** | |  |
|  | **Exposure** | **β Coefficient** | **p-value** | **β Coefficient** | **p-value** | **N** |
|  |  | **(95% CI)** |  | **(95% CI)** |  |  |
| **No. of unique clinically relevant ARGs** | **Animal Exposure** | | |  |  |  |
|  | Upper tertile | Reference |  | Reference |  | 4 |
|  | Middle tertile | -4.2 (-10.52, 2.12) | 0.20 | -5.37 (-11.43, 0.68) | 0.09 | 10 |
|  | Lower tertile | -5.36 (-11.41, 0.7) | 0.09 | -5.58 (-11.46, 0.29) | 0.07 | 14 |
|  | Zero | -0.3 (-6.05, 5.45) | 0.92 | -2.08 (-7.72, 3.56) | 0.47 | 25 |
|  | **Sanitation & DW** | | |  |  |  |
|  | Other | Reference |  | Reference |  | 21 |
|  | CS&PW | -4.26 (-7.35, -1.17) | 0.01 | -3.52 (-6.74, -0.30) | 0.04 | 30 |
|  | **Piped Water** |  |  |  |  |  |
|  | Not piped | Reference |  | Reference |  | 14 |
|  | 0-6 days | -3.26 (-6.96, 0.43) | 0.09 | -1.51 (-5.72, 2.70) | 0.49 | 21 |
|  | 7 days | -5.63 (-9.45, -1.82) | 0.01 | -3.45 (-8.20, 1.29) | 0.16 | 18 |
| **Nonpareil Sequence Diversity** | **Animal Exposure** | |  |  |  |  |
|  | Upper tertile | Reference |  | Reference |  | 4 |
|  | Middle tertile | -0.94 (-1.62, -0.27) | 0.01 | -1.03 (-1.72, -0.35) | <0.001 | 10 |
|  | Lower tertile | -1.18 (-1.83, -0.54) | <0.001 | -1.29 (-1.96, -0.63) | <0.001 | 14 |
|  | Zero | -0.93 (-1.54, -0.31) | <0.001 | -1.09 (-1.73, -0.45) | <0.001 | 25 |
|  | **Sanitation & DW** | |  |  |  |  |
|  | Other | Reference |  | Reference |  | 21 |
|  | CS&PW | -0.08 (-0.45, 0.28) | 0.66 | 0.02 (-0.39, 0.43) | 0.92 | 30 |
|  | **Piped Water** |  |  |  |  |  |
|  | Not piped | Reference |  | Reference |  | 14 |
|  | 0-6 days | 0.06 (-0.36, 0.49) | 0.77 | 0.29 (-0.21, 0.79) | 0.26 | 21 |
|  | 7 days | -0.32 (-0.75, 0.12) | 0.16 | -0.24 (-0.8, 0.32) | 0.41 | 18 |
| **Clinically Relevant ARG Abundance** | **Animal Exposure** | |  |  |  |  |
|  | Upper tertile | Reference |  | Reference |  | 4 |
|  | Middle tertile | 0.01 (-0.06, 0.08) | 0.78 | 0 (-0.07, 0.07) | 0.98 | 10 |
|  | Lower tertile | -0.03 (-0.1, 0.03) | 0.33 | -0.04 (-0.1, 0.03) | 0.26 | 14 |
|  | Zero | 0 (-0.06, 0.06) | 0.97 | -0.02 (-0.08, 0.05) | 0.63 | 25 |
|  | **Sanitation & DW** | |  |  |  |  |
|  | Other | Reference |  | Reference |  | 21 |
|  | CS&PW | -0.03 (-0.06, 0.01) | 0.14 | -0.02 (-0.05, 0.02) | 0.32 | 30 |
|  | **Piped Water** |  |  |  |  |  |
|  | Not piped | Reference |  | Reference |  | 14 |
|  | 0-6 days | -0.06 (-0.09, -0.02) | 0.004 | -0.05 (-0.1, -0.01) | 0.02 | 21 |
|  | 7 days | -0.06 (-0.09, -0.02) | 0.005 | -0.04 (-0.09, 0.00) | 0.07 | 18 |
| **% *E. coli* Relative Abundance** | **Animal Exposure** | |  |  |  |  |
|  | Upper tertile | Reference |  | Reference |  | 4 |
|  | Middle tertile | -0.24 (-0.8, 0.33) | 0.41 | -0.26 (-0.82, 0.30) | 0.37 | 9 |
|  | Lower tertile | -0.24 (-0.77, 0.29) | 0.38 | -0.32 (-0.87, 0.22) | 0.25 | 14 |
|  | Zero | 0.12 (-0.39, 0.62) | 0.65 | 0.03 (-0.5, 0.55) | 0.92 | 25 |
|  | **Sanitation & DW** | |  |  |  |  |
|  | Other | Reference |  | Reference |  | 21 |
|  | CS&PW | -0.3 (-0.57, -0.03) | 0.04 | -0.23 (-0.53, 0.07) | 0.14 | 29 |
|  | **Piped Water** |  |  |  |  |  |
|  | Not piped | Reference |  | Reference |  | 13 |
|  | 0-6 days | -0.07 (-0.41, 0.27) | 0.70 | 0.11 (-0.29, 0.51) | 0.59 | 21 |
|  | 7 days | -0.29 (-0.64, 0.06) | 0.11 | -0.12 (-0.56, 0.32) | 0.61 | 18 |
| **% *K. pneumoniae* Relative Abundance** | **Animal Exposure** | |  |  |  |  |
|  | Upper tertile | Reference |  | Reference |  | 4 |
|  | Middle tertile | 0 (-0.64, 0.63) | 0.99 | -0.05 (-0.72, 0.62) | 0.89 | 10 |
|  | Lower tertile | 0 (-0.61, 0.61) | 0.99 | -0.03 (-0.69, 0.62) | 0.92 | 14 |
|  | Zero | 0.3 (-0.28, 0.88) | 0.32 | 0.26 (-0.37, 0.89) | 0.42 | 25 |
|  | **Sanitation & DW** | |  |  |  |  |
|  | Other | Reference |  | Reference |  | 21 |
|  | CS&PW | -0.04 (-0.36, 0.28) | 0.81 | 0 (-0.36, 0.36) | 0.99 | 30 |
|  | **Piped Water** |  |  |  |  |  |
|  | Not piped | Reference |  | Reference |  | 14 |
|  | 0-6 days | -0.24 (-0.62, 0.14) | 0.22 | -0.4 (-0.85, 0.04) | 0.08 | 21 |
|  | 7 days | -0.15 (-0.54, 0.24) | 0.44 | -0.11 (-0.61, 0.39) | 0.67 | 18 |

| **Table S4:** Linear model results for the microbiological outcomes with animal exposure, DW & San and piped water availability in children. | | | | | | |
| --- | --- | --- | --- | --- | --- | --- |
|  |  | **Unadjusted** | | **Adjusted** | | |
|  | **Exposure** | **β Coefficient** | **p-value** | **β Coefficient** | **p-value** | **N** |
|  |  | **(95% CI)** |  | **(95% CI)** |  |  |
| **No. of unique clinically relevant ARGs** | **Animal Exposure** | |  |  |  |  |
|  | Upper tertile | Reference |  | Reference |  | 29 |
|  | Middle tertile | 7.87 (0, 15.73) | 0.05 | 5.24 (-2.58, 13.06) | 0.19 | 31 |
|  | Lower tertile | 11.61 (3.1, 20.12) | 0.01 | 7.85 (-0.33, 16.03) | 0.06 | 32 |
|  | Zero | 5.44 (0.12, 10.77) | 0.05 | 1.57 (-4.66, 7.8) | 0.62 | 58 |
|  | **Sanitation & DW** | |  |  |  |  |
|  | Other | Reference |  | Reference |  | 47 |
|  | CS&PW | -0.45 (-5.99, 5.09) | 0.87 | -5.42 (-11.64, 0.79) | 0.09 | 103 |
|  | **Piped Water** |  |  |  |  |  |
|  | Not piped | Reference |  | Reference |  | 32 |
|  | 0-6 days | 5.84 (0.15, 11.53) | 0.04 | 4.72 (-2.09, 11.53) | 0.17 | 28 |
|  | 7 days | 3.02 (-3.51, 9.55) | 0.36 | 0.99 (-7.88, 9.86) | 0.83 | 19 |
| **Nonpareil Sequence Diversity** | **Animal Exposure** | |  |  |  |  |
|  | Upper tertile | Reference |  | Reference |  | 29 |
|  | Middle tertile | -0.33 (-0.83, 0.18) | 0.21 | 0.08 (-0.35, 0.52) | 0.70 | 31 |
|  | Lower tertile | -0.63 (-1.16, -0.1) | 0.02 | 0.21 (-0.28, 0.7) | 0.40 | 32 |
|  | Zero | -0.74 (-1.26, -0.22) | 0.01 | 0.05 (-0.41, 0.5) | 0.84 | 58 |
|  | **Sanitation & DW** | |  |  |  |  |
|  | Other | Reference |  | Reference |  | 47 |
|  | CS&PW | -0.42 (-0.71, -0.12) | 0.01 | -0.1 (-0.37, 0.17) | 0.47 | 103 |
|  | **Piped Water** |  |  |  |  |  |
|  | Not piped | Reference |  | Reference |  | 32 |
|  | 0-6 days | -0.15 (-0.63, 0.33) | 0.55 | -0.29 (-0.75, 0.17) | 0.22 | 28 |
|  | 7 days | -0.56 (-1.1, -0.02) | 0.04 | -0.64 (-1.14, -0.15) | 0.01 | 19 |
| **Clinically Relevant ARG Abundance** | **Animal Exposure** | |  |  |  |  |
|  | Upper tertile | Reference |  | Reference |  | 29 |
|  | Middle tertile | 0.12 (-0.03, 0.26) | 0.11 | 0.1 (-0.04, 0.23) | 0.15 | 31 |
|  | Lower tertile | 0.09 (-0.02, 0.21) | 0.12 | 0.05 (-0.07, 0.17) | 0.40 | 32 |
|  | Zero | 0.17 (0.03, 0.31) | 0.02 | 0.11 (-0.06, 0.27) | 0.21 | 58 |
|  | **Sanitation & DW** | |  |  |  |  |
|  | Other | Reference |  | Reference |  | 47 |
|  | CS&PW | -0.07 (-0.16, 0.03) | 0.18 | -0.09 (-0.2, 0.01) | 0.09 | 103 |
|  | **Piped Water** |  |  |  |  |  |
|  | Not piped | Reference |  | Reference |  | 32 |
|  | 0-6 days | 0.16 (-0.01, 0.33) | 0.06 | 0.12 (-0.03, 0.28) | 0.12 | 28 |
|  | 7 days | 0.05 (-0.03, 0.14) | 0.23 | 0.03 (-0.07, 0.12) | 0.60 | 19 |
| **% *E. coli* Relative Abundance** | **Animal Exposure** | |  |  |  |  |
|  | Upper tertile | Reference |  | Reference |  | 29 |
|  | Middle tertile | 3.37 (-0.81, 7.56) | 0.11 | 2.54 (-1.29, 6.36) | 0.19 | 31 |
|  | Lower tertile | 0.17 (-2.51, 2.85) | 0.90 | -1.42 (-4.38, 1.54) | 0.35 | 32 |
|  | Zero | 3.06 (-0.01, 6.13) | 0.05 | 0.97 (-2.39, 4.32) | 0.57 | 58 |
|  | **Sanitation & DW** | |  |  |  |  |
|  | Other | Reference |  | Reference |  | 47 |
|  | CS&PW | -2.1 (-4.56, 0.36) | 0.09 | -3.37 (-6.49, -0.24) | 0.03 | 103 |
|  | **Piped Water** | |  |  |  |  |
|  | Not piped | Reference |  | Reference |  | 32 |
|  | 0-6 days | 2.46 (-0.17, 5.09) | 0.07 | 1.36 (-1.11, 3.83) | 0.28 | 28 |
|  | 7 days | 1.27 (-0.7, 3.24) | 0.21 | 0.5 (-1.56, 2.55) | 0.63 | 19 |
| **% *K. pneumoniae* Relative Abundance** | **Animal Exposure** | |  |  |  |  |
|  | Upper tertile | Reference |  | Reference |  | 29 |
|  | Middle tertile | 0.59 (-0.77, 1.95) | 0.39 | -0.31 (-2.5, 1.87) | 0.78 | 31 |
|  | Lower tertile | 3.03 (-0.74, 6.8) | 0.12 | 1.69 (-1.17, 4.55) | 0.25 | 32 |
|  | Zero | 2.42 (0.03, 4.82) | 0.05 | 0.63 (-2.11, 3.36) | 0.65 | 58 |
|  | **Sanitation & DW** | |  |  |  |  |
|  | Other | Reference |  | Reference |  | 47 |
|  | CS&PW | 3.03 (-0.16, 6.21) | 0.06 | 1.91 (-1.74, 5.56) | 0.31 | 103 |
|  | **Piped Water** |  |  |  |  |  |
|  | Not piped | Reference |  | Reference |  | 32 |
|  | 0-6 days | 1.17 (-0.18, 2.52) | 0.09 | 1.25 (-0.48, 2.98) | 0.16 | 28 |
|  | 7 days | -0.96 (-2.44, 0.53) | 0.21 | -1.83 (-4.96, 1.3) | 0.25 | 19 |

| **Table S5:** Linear model results for animal exposure in subgroups (mothers). | | | | | | | | |
| --- | --- | --- | --- | --- | --- | --- | --- | --- |
|  |  |  | **Unadjusted** | | **Adjusted** | |  | |
|  | **Sub**  **group** | **Exposure** | **β Coefficient (95% CI)** | **p-value** | **β Coefficient**  **(95% CI)** | **p-value** | **Diff. Models** | **N** |
|  |  |  |  |  |  |  | **p-value** |  |
| **No. of unique clinically relevant ARGs** | SC & PW | Upper tertile | Reference |  | Reference |  | 0.24 | 2 |
|  |  | Middle tertile | -1.83 (-8.85, 5.19) | 0.62 | -2.10 (-10.50, 6.29) | 0.63 |  | 6 |
|  |  | Lower Tertile | -5.67 (-12.69,1.35) | 0.13 | -5.42 (-13.56, 2.73) | 0.21 |  | 6 |
|  |  | Zero | 2.14 (-4.75, 9.04) | 0.55 | 1.69 (-6.80, 10.19) | 0.70 |  | 7 |
|  | Other | Upper tertile | Reference |  | Reference |  |  | 2 |
|  |  | Middle tertile | -6.0 (-15.9, 3.92) | 0.25 | -11.57 (-21.09, -2.04) | 0.03 |  | 4 |
|  |  | Lower Tertile | -6.0 (-15.1, 3.05) | 0.21 | -10.31 (-19.29, -1.34) | 0.03 |  | 8 |
|  |  | Zero | -3.31 (-11.9, 5.28) | 0.46 | -6.31 (-14.36, 1.75) | 0.14 |  | 16 |
| **Nonpareil Sequence Diversity** | SC & PW | Upper tertile | Reference |  | Reference |  | 0.34 | 2 |
|  |  | Middle tertile | -1.05 (-2.12, 0.01) | 0.07 | -1.16 (-2.42, 0.11) | 0.09 |  | 6 |
|  |  | Lower Tertile | -1.75 (-2.82, -0.69) | <0.001 | -1.85 (-3.08, -0.63) | 0.01 |  | 6 |
|  |  | Zero | -1.13 (-2.18, -0.09) | 0.05 | -1.2 (-2.48, 0.07) | 0.09 |  | 7 |
|  | Other | Upper tertile | Reference |  | Reference |  |  | 2 |
|  |  | Middle tertile | -0.89 (-1.79, 0.02) | 0.07 | -1.19 (-2.13, -0.24) | 0.02 |  | 4 |
|  |  | Lower Tertile | -0.7 (-1.53, 0.12) | 0.11 | -0.91 (-1.8, -0.02) | 0.06 |  | 8 |
|  |  | Zero | -0.68 (-1.46, 0.1) | 0.10 | -0.95 (-1.75, -0.16) | 0.03 |  | 16 |
| **Clinically Relevant ARG Abundance** | SC & PW | Upper tertile | Reference |  | Reference |  | 0.38 | 2 |
|  |  | Middle tertile | -0.02 (-0.07, 0.03) | 0.38 | -0.02 (-0.07, 0.04) | 0.52 |  | 6 |
|  |  | Lower Tertile | -0.04 (-0.08, 0.01) | 0.16 | -0.03 (-0.08, 0.02) | 0.29 |  | 6 |
|  |  | Zero | 0 (-0.04, 0.05) | 0.87 | 0.01 (-0.05, 0.06) | 0.75 |  | 7 |
|  | Other | Upper tertile | Reference |  | Reference |  |  | 2 |
|  |  | Middle tertile | 0.06 (-0.06, 0.18) | 0.34 | 0.01 (-0.12, 0.14) | 0.87 |  | 4 |
|  |  | Lower Tertile | -0.03 (-0.14, 0.08) | 0.57 | -0.07 (-0.19, 0.05) | 0.26 |  | 8 |
|  |  | Zero | 0 (-0.11, 0.1) | 0.95 | -0.03 (-0.14, 0.08) | 0.57 |  | 16 |
| **% *E. coli* Relative Abundance** | SC & PW | Upper tertile | Reference |  | Reference |  |  | 2 |
|  |  | Middle tertile | -0.32 (-0.63, -0.02) | 0.06 | -0.29 (-0.63, 0.06) | 0.13 | 0.78 | 6 |
|  |  | Lower Tertile | -0.43 (-0.74, -0.13) | 0.01 | -0.46 (-0.8, -0.13) | 0.02 |  | 6 |
|  |  | Zero | -0.01 (-0.31, 0.3) | 0.97 | 0.09 (-0.26, 0.44) | 0.63 |  | 7 |
|  | Other | Upper tertile | Reference |  | Reference |  |  | 2 |
|  |  | Middle tertile | -0.07 (-1.16, 1.02) | 0.90 | -0.27 (-1.47, 0.93) | 0.66 |  | 4 |
|  |  | Lower Tertile | -0.09 (-1.03, 0.85) | 0.85 | -0.36 (-1.47, 0.75) | 0.53 |  | 8 |
|  |  | Zero | 0.22 (-0.68, 1.11) | 0.64 | 0.00 (-0.99, 1.00) | 1 |  | 16 |
| **% *K. pneumoniae* Relative Abundance** | SC & PW | Upper tertile | Reference |  | Reference |  |  | 2 |
|  |  | Middle tertile | -0.02 (-0.54, 0.51) | 0.95 | -0.04 (-0.64, 0.55) | 0.89 | 1 | 6 |
|  |  | Lower Tertile | -0.04 (-0.57, 0.49) | 0.88 | -0.08 (-0.66, 0.49) | 0.78 |  | 6 |
|  |  | Zero | 0.32 (-0.2, 0.84) | 0.24 | 0.39 (-0.21, 0.99) | 0.22 |  | 7 |
|  | Other | Upper tertile | Reference |  | Reference |  |  | 2 |
|  |  | Middle tertile | 0 (-1.2, 1.2) | 1 | -0.39 (-1.76, 0.99) | 0.59 |  | 4 |
|  |  | Lower Tertile | 0.03 (-1.06, 1.12) | 0.95 | -0.18 (-1.48, 1.11) | 0.79 |  | 8 |
|  |  | Zero | 0.32 (-0.71, 1.36) | 0.55 | 0.12 (-1.04, 1.28) | 0.84 |  | 16 |

| **Table S6:** Linear model results for animal exposure in subgroups (children). | | | | | | | | | | | | | |
| --- | --- | --- | --- | --- | --- | --- | --- | --- | --- | --- | --- | --- | --- |
|  |  | |  | | **Unadjusted** | | | **Adjusted** | | | |  |  |
|  | **Sub**  **group** | | **Exposure** | | **β Coefficient (95% CI)** | **p-value** | | **β Coefficient**  **(95% CI)** | | **p-value** | | **Diff. Models**  **p-value** | **N** |
| **No. of unique clinically relevant ARGs** | | SC & PW | Upper tertile | Reference | | |  | | Reference | |  | 0.001 | 3 |
|  |  |  | Middle tertile | 23.51 (13.38, 33.6) | | | <0.001 | | 19.95 (-1.34, 41.23) | | 0.07 |  | 10 |
|  |  |  | Lower Tertile | 18.08 (7.28, 28.9) | | | <0.001 | | 13.57 (-4.07, 31.21) | | 0.13 |  | 15 |
|  |  |  | Zero | 9.65 (2.11, 17.2) | | | 0.01 | | 9.74 (-4.21, 23.7) | | 0.17 |  | 18 |
|  |  | Other | Upper tertile | Reference | | |  | | Reference | |  |  | 26 |
|  |  |  | Middle tertile | 2.99 (-5.24, 11.2) | | | 0.48 | | 1.27 (-6.71, 9.25) | | 0.76 |  | 21 |
|  |  |  | Lower Tertile | 9.32 (-3.14, 21.8) | | | 0.14 | | 7.01 (-5.58, 19.6) | | 0.28 |  | 16 |
|  |  |  | Zero | 6.23 (0.05, 12.4) | | | 0.05 | | 3.48 (-4.22, 11.18) | | 0.38 |  | 38 |
| **Nonpareil Sequence Diversity** | | SC & PW | Upper tertile | Reference | | |  | | Reference | |  | <0.001 | 3 |
|  |  |  | Middle tertile | 0.85 (-0.1, 1.8) | | | 0.08 | | 1.16 (0.61, 1.72) | | <0.001 |  | 10 |
|  |  |  | Lower Tertile | 0.82 (-0.09, 1.72) | | | 0.08 | | 1.13 (0.57, 1.68) | | <0.001 |  | 15 |
|  |  |  | Zero | 0.72 (-0.15, 1.58) | | | 0.10 | | 0.78 (0.27, 1.28) | | <0.001 |  | 18 |
|  |  | Other | Upper tertile | Reference | | |  | | Reference | |  |  | 26 |
|  |  |  | Middle tertile | -0.31 (-0.84, 0.22) | | | 0.25 | | -0.13 (-0.65, 0.39) | | 0.63 |  | 21 |
|  |  |  | Lower Tertile | -0.76 (-1.41, -0.12) | | | 0.02 | | 0.23 (-0.31, 0.77) | | 0.40 |  | 16 |
|  |  |  | Zero | -0.91 (-1.48, -0.34) | | | <0.001 | | 0.1 (-0.49, 0.69) | | 0.74 |  | 38 |
| **Clinically Relevant ARG Abundance** | | SC & PW | Upper tertile | Reference | | |  | | Reference | |  | 0.10 | 3 |
|  |  |  | Middle tertile | 0.09 (-0.04, 0.22) | | | 0.19 | | 0.03 (-0.11, 0.16) | | 0.71 |  | 10 |
|  |  |  | Lower Tertile | 0.07 (-0.09, 0.23) | | | 0.41 | | 0.02 (-0.13, 0.16) | | 0.82 |  | 15 |
|  |  |  | Zero | 0.06 (-0.05, 0.16) | | | 0.28 | | 0.06 (-0.06, 0.18) | | 0.30 |  | 18 |
|  |  | Other | Upper tertile | Reference | | |  | | Reference | |  |  | 26 |
|  |  |  | Middle tertile | 0.13 (-0.06, 0.32) | | | 0.16 | | 0.17 (-0.01, 0.36) | | 0.07 |  | 21 |
|  |  |  | Lower Tertile | 0.13 (-0.03, 0.28) | | | 0.10 | | 0.1 (-0.09, 0.29) | | 0.30 |  | 16 |
|  |  |  | Zero | 0.24 (0.04, 0.44) | | | 0.02 | | 0.19 (-0.07, 0.45) | | 0.16 |  | 38 |
| **% *E. coli* Relative Abundance** | | SC & PW | Upper tertile | Reference | | |  | | Reference | |  |  | 3 |
|  |  |  | Middle tertile | 6.9 (2.97, 10.82) | | | <0.001 | | 6.73 (2.62, 10.84) | | <0.001 | 0.09 | 10 |
|  |  |  | Lower Tertile | 1.37 (-0.51, 3.26) | | | 0.15 | | 0.74 (-2.53, 4.01) | | 0.66 |  | 15 |
|  |  |  | Zero | 2.79 (0.13, 5.46) | | | 0.04 | | 2.73 (-1.35, 6.81) | | 0.19 |  | 18 |
|  |  | Other | Upper tertile | Reference | | |  | | Reference | |  |  | 26 |
|  |  |  | Middle tertile | 2.73 (-2.83, 8.28) | | | 0.34 | | 2.52 (-2.14, 7.18) | | 0.29 |  | 21 |
|  |  |  | Lower Tertile | 1.68 (-1.56, 4.93) | | | 0.31 | | -1.74 (-6.29, 2.81) | | 0.45 |  | 16 |
|  |  |  | Zero | 4.25 (0.26, 8.24) | | | 0.04 | | 0.51 (-4.79, 5.81) | | 0.85 |  | 38 |
| **% *K. pneumoniae* Relative Abundance** | | SC & PW | Upper tertile | Reference | | |  | | Reference | |  |  | 3 |
|  |  |  | Middle tertile | 2.33 (0.12, 4.54) | | | 0.04 | | 2.31 (-7.01, 11.63) | | 0.63 | 0.06 | 10 |
|  |  |  | Lower Tertile | 5.62 (-0.58, 11.82) | | | 0.08 | | 6.91 (-1.27, 15.08) | | 0.10 |  | 15 |
|  |  |  | Zero | 4.37 (0, 8.74) | | | 0.05 | | 6.86 (-1.04, 14.76) | | 0.09 |  | 18 |
|  |  | Other | Upper tertile | Reference | | |  | | Reference | |  |  | 26 |
|  |  |  | Middle tertile | -1.03 (-2.13, 0.07) | | | 0.07 | | -1.27 (-2.44, -0.09) | | 0.03 |  | 21 |
|  |  |  | Lower Tertile | 0.46 (-2.33, 3.25) | | | 0.75 | | -0.17 (-1.61, 1.27) | | 0.82 |  | 16 |
|  |  |  | Zero | 1.44 (-0.81, 3.7) | | | 0.21 | | 0.29 (-1.33, 1.91) | | 0.73 |  | 38 |
